# Attention-deficit/hyperactive disorder pre-adulthood and later adverse health outcomes: The 1987 Finnish birth cohort study

**DOI:** 10.1101/2025.06.24.25330197

**Authors:** G. David Batty, Mika Gissler, Seyed Ehsan Mousavi, Varun Warrier, Tamsin Ford, Markus Keski-Säntti

## Abstract

Whereas attention-deficit/hyperactive disorder (ADHD) is correlated with later risk of depression, anxiety, and substance misuse, the relationship with other health endpoints is uncertain. In a full-nation birth cohort study, we used a phenotype-wide approach to explore the influence of an ADHD diagnosis in childhood/adolescence with later disease and injury. Comprising 53147 (25731 female) children born in a single year, the 1987 Finnish Birth Cohort was generated from linkage of routinely collected data. Using international classification disease codes, ADHD diagnosis was captured from in- and out-patient hospital records up to age 18 years and study members continued to be surveilled for other diagnoses until 2020 (aged 33 years). In logistic regression analyses, effect estimates were adjusted for education achievement, family socioeconomic status, and multiple comparisons. Pre-adulthood, 0.43% (N=228) of study members were diagnosed with ADHD. In people with ADHD relative to population controls, there was a heightened risk of developing all 17 specific health endpoints examined. Of these, only 5 reached statistical significance after correction for socioeconomic status, education, and multiple comparison (odds ratio; 99.7%): substance abuse disorders (2.27; 1.28, 3.81), mood disorders (2.46; 1.50, 3.90), neurotic disorders (2.12; 1.25, 3.43), epilepsy (4.65; 1.86, 9.75), and poisoning (2.30; 1.02, 4.51). In the present study, children and adolescents with ADHD had an increased future burden of psychological and neurological conditions but not somatic disorders.

## Introduction

First posited over four decades ago, the developmental origins of adult disease – the notion that adverse health in later life is influenced by early life exposures – has traditionally focused on the deterministic role of markers of *physical* development,^1^ such as body weight from birth through to adolescence, physical stature, and age at onset of puberty.^2–5^ By contrast, it is only in the last 5 years that the role of *neurological* development and associated disorders such as intellectual disability, autism, Tourette syndrome/Tic disorder, and attention-deficit/hyperactive disorder (ADHD) in the occurrence of adult disease and injury have been systematically examined.^6–9^ Of these neurodevelopmental conditions, if ADHD is shown to be connected to later health, it holds the largest potential for event prevention given that it is the most commonly occurring neurodevelopmental disorder (3-7% prevalence^10^), and its core symptoms of age-inappropriate levels of distraction, restlessness, and impulsivity are eminently modifiable via prescription psychostimulants and psychosocial intervention.^11^

The greater prevalence of obesity, unfavourable health behaviours such as cigarette smoking and heavy alcohol intake, educational challenges, and poverty in people with ADHD relative to population controls^12^ suggests the potential development of an array of related health outcomes in affected individuals. Thus, people with people with this neurodevelopmental disorder appear to have a greater burden of psychological ill-health and addiction such as depression,^13,14^ anxiety,^15^ and substance misuse,^16^ although these are not universal observations.^17^ Most recently, nascent findings from extended follow-up in cohort studies generated from linked administrative records indicate that younger people diagnosed with ADHD subsequently experience around twice the rate of somatic disease, including diabetes,^18^ dementia,^19^ and cardiovascular disease.^20^

Owing to the core symptoms of ADHD, there are also reasons to anticipate associations with lesser-examined health endpoints such as respiratory illness and unintentional injury. Thus, potentially via a lack of compliance with preventative measures such as mask wearing and handwashing, ADHD-affected individuals have been shown to have a higher burden of COVID-19^21^ compared with rates in the general population; the connection with other common pulmonary infections, such as influenza and pneumonia, is untested however. Unintentional injury, such as road traffic accidents^22^ may also be more common in people with this neurodevelopmental disorder because of inattention, impulsivity, and longer reaction time.^23^

To address the general paucity of evidence for health outcomes in relation to antecedent ADHD, and to test some of these hypothesised associations, we undertook analyses of an on-going, full nation birth cohort of individuals born in Finland in 1987 with routinely collected data on ADHD, covariates, and episodes of adverse health. Our analyses also allow us to examine the potential explanatory role of confounding factors, a perennial concern in observational studies. In the present context, these include socioeconomic background and educational performance, both of which have been consistently linked to ADHD^24^ and a series of health conditions.^25^

## Methods

Described in detail elsewhere,^26^ the 1987 Finnish Birth Cohort was generated from the linkage of routinely collected data using the personal identification number allocated to every citizen. The cohort comprises 59476 people (figure 1). Study approval was provided by the Ethical committee at the Finnish Institute for Health and Welfare (§28/2009). The composition of this manuscript conforms to the Strengthening the Reporting of Observational Studies in Epidemiology (STROBE) Statement.^27^

**Figure 1.**
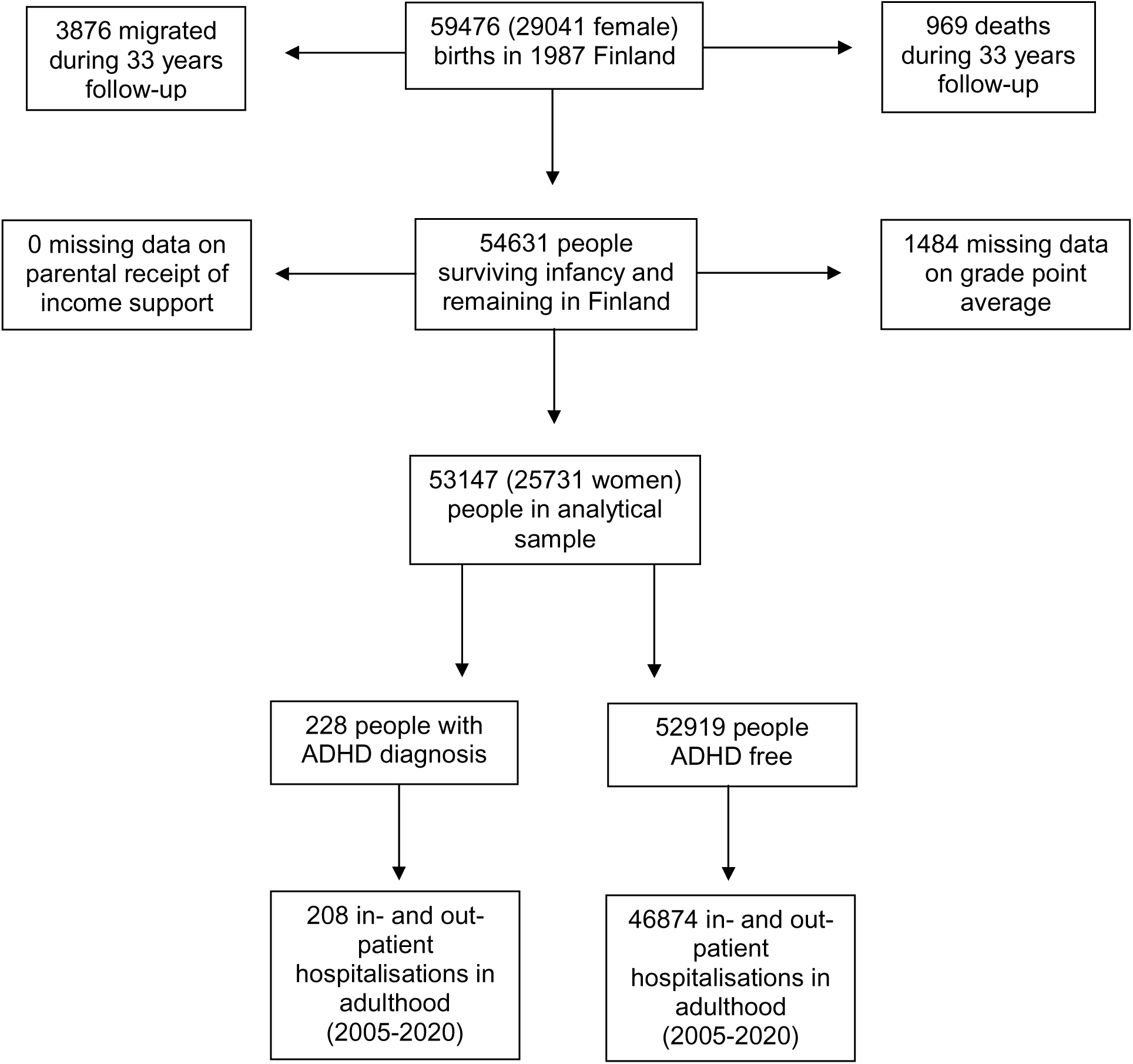
Flow of study members into the analytical sample

### Ascertainment of ADHD and health outcomes

The Finnish Care Register for Health Care (formerly the Hospital Discharge Register) captures in-(1969-present) and out-patient care episodes (1998-present) from all hospitals in the country.^28^ Coded according to the International Classification of Diseases (ICD) versions 9 (1987-1995) and 10 (1996-2004), a diagnosis of ADHD was denoted by codes 314 (ICD-9) and F90 (ICD-10). ICD codes for the health outcomes in the present analyses are listed in figures 2 and 3.

**Figure 2.**
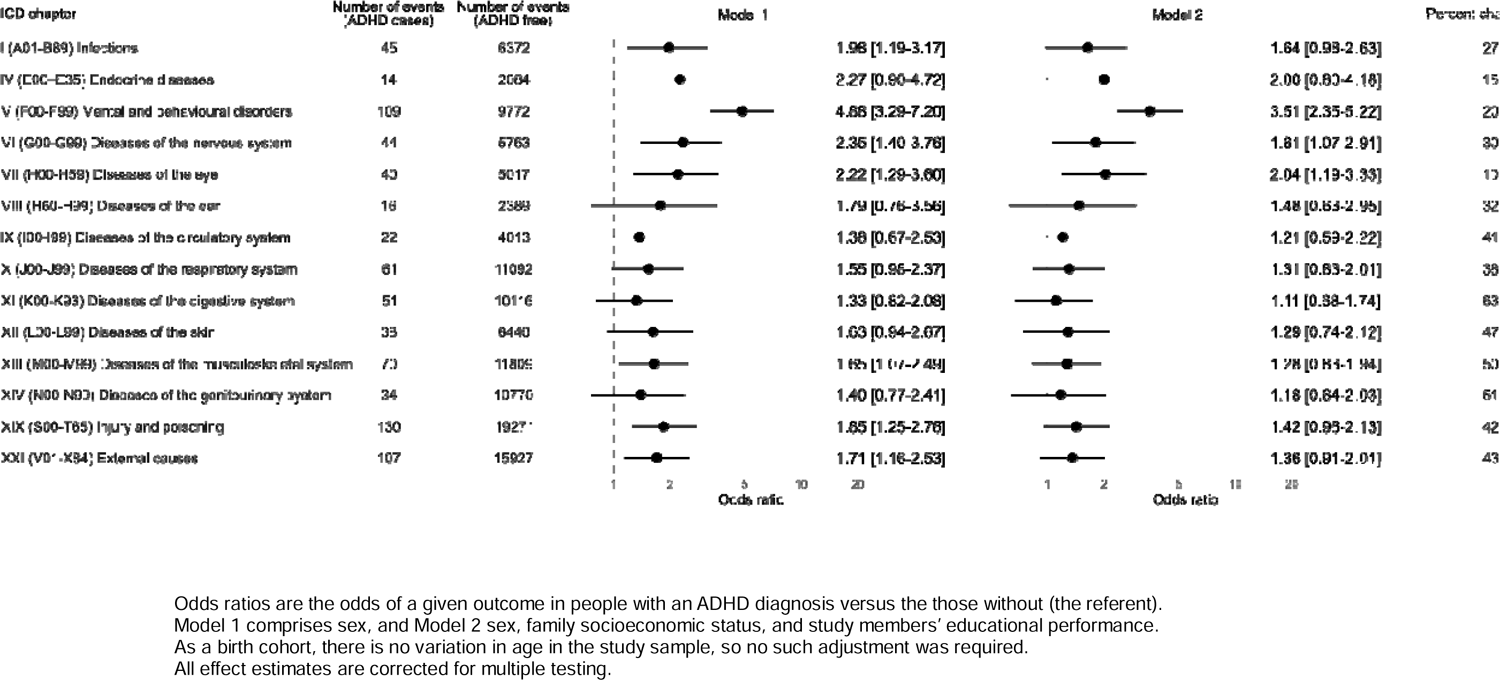
Odds ratios (99.6% confidence intervals) for the association between ADHD diagnosis and subsequent hospitalisation for 14 health conditions (N=53147)

### Assessment of covariates

Parental socioeconomic status, extracted from the Register of Social Assistance, was denoted by the duration (months) the study members’ carer, typically the parent, was in receipt of government income support. Longer duration denotes greater socioeconomic disadvantage. For educational performance of the study member, we used school grade point average (score range: 4-10) as provided by the Finnish National Agency for Education. A greater score is indicative of higher educational performance.

### Derivation of the analytical sample and statistical analyses

From the 59476 births (29041 females) in 1987, we excluded people who subsequently died or emigrated before the end of follow-up 33 years later (31 December 2020) and those missing covariate data (figure 1). Analyses were based on a sample of 53147 individuals (25731 women) in which ADHD on or before the 18^th^ birthday of the study members (calendar year 2005) was related to the occurrence of adverse health events between age 19 and 33 (2005-2020). During follow-up, a study member could conceivably not have diagnosis of a health episode, a single diagnosis, or multiple diagnoses made on single or multiple occasions. When there were multiple events recorded, each contributed separately to the analyses of a specific health outcome. Thus, in preliminary analyses, of the 228 individuals with ADHD diagnosis, during 15 years of follow-up, 208 had one or more contact with hospital services for a total of 10494 contacts. In the ADHD-free group, of the 52919 people, 46874 had contact with hospital services for a total of 1452935 contacts.

We used logistic regression analyses to computed odds ratios to summarise the relationship between ADHD and later event diagnosis. Taking a phenotype-wide approach, those outcomes with 10 or more events in the ADHD group and population controls qualified for inclusion in our analyses. In minimally-adjusted analyses (model 1), we controlled odds ratios for sex; with this being a single-year birth cohort, no adjustment for age was required. Further adjustment was then made for the educational performance of the study member and family socioeconomic status (model 2). Using the Bonferroni correction for multiple testing was applied to the computation of all effect estimates, with statistical significance set at p=0.004 (96.6% confidence intervals) for the analyses featured in figure 2 on ICD chapters (14 outcomes) and p=0.003 (99.7 confidence intervals) for the analyses featured in figure 3 for chapter disaggregation (17 outcomes).^29^ Change in odds ratios following adjustment for multiple covariates relative to minimal correction was computed using standard formulae ((ln odds ratio[Model 1] – ln odds ratio [Model 2]) / ln odds ratio[Model 1]) with the product expressed as a percentage.^30^

**Figure 3.**
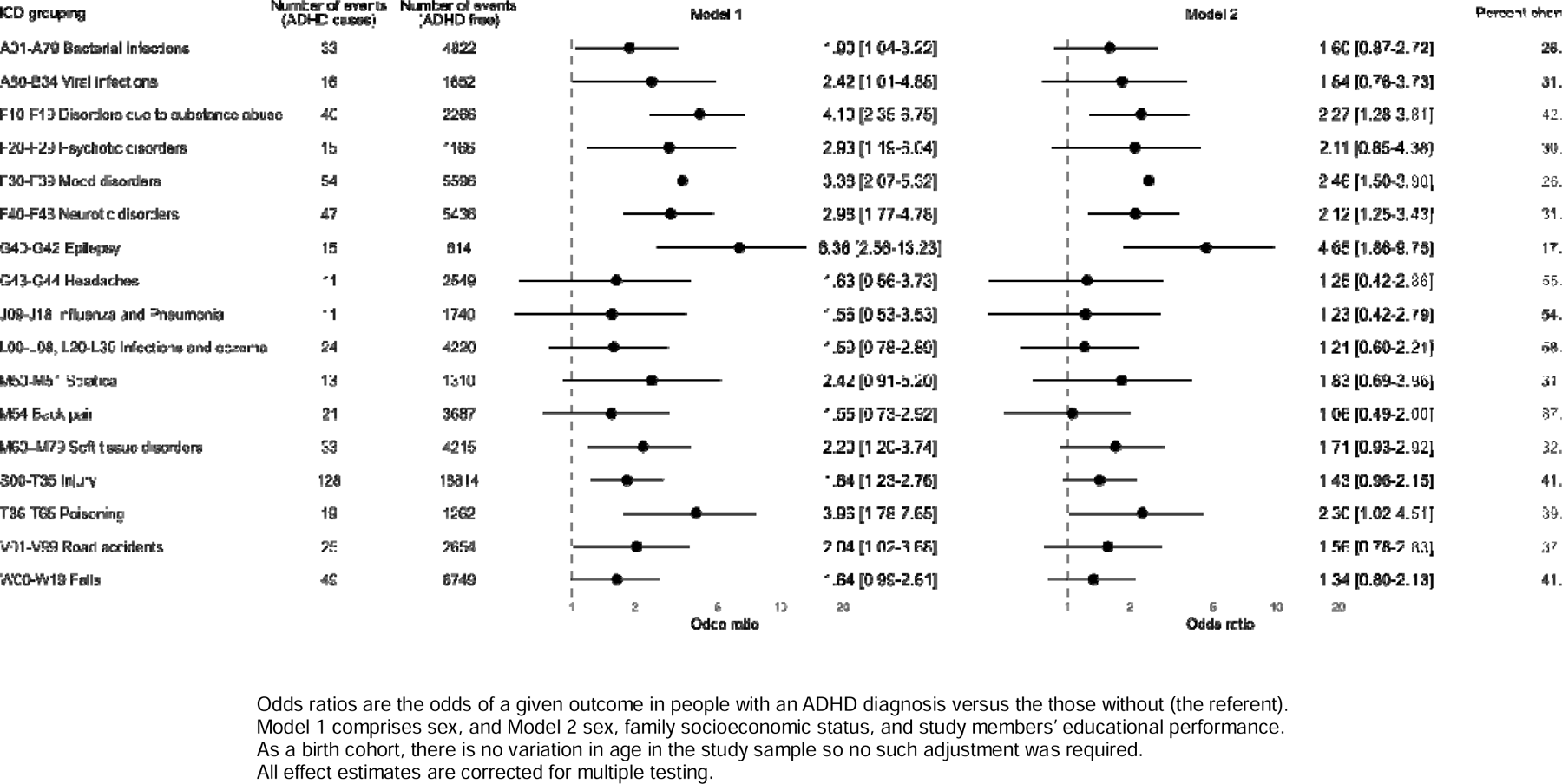
Odds ratios (99.7% confidence intervals) for the association between ADHD diagnosis and subsequent hospitalisation for 17 health conditions (N=53147)

## Results

In the analytical sample of 53147 individuals (25731 women), 0.43% (N=228) were diagnosed with ADHD on or before their 18^th^ birthday. Relative to their unaffected counterparts, study members with ADHD were more likely to be male (51 versus 86%), and have a less favourable cluster of social characteristics. Thus, people with ADHD were more likely to have a lower grade point average (6.9 versus 7.8) and have parents who were in receipt of state welfare payments for a longer period (48.8 versus 17.6 months).

After fifteen years of follow-up, 89% of the full cohort (N=47082) had experienced in- or out-patient contact with a hospital for any health event (figure 1 and table 1), with hospital visits marginally more common in people with ADHD (91%) than their unaffected counterparts (89%). In figure 2, we depict the association between ADHD and subsequent hospitalisation for outcomes denoted by 14 ICD chapters. In analyses corrected for sex and multiple comparisons (model 1), there was a higher risk of all 14 endpoints in people diagnosed with ADHD relative to population controls, although statistical significance was only apparent for half. Where the confidence interval did not include unity, the magnitude of the odds ratio (99.6% confidence interval) ranged between 1.65 (1.07, 2.49) for diseases of the musculoskeletal system and 4.88 (3.29, 7.20) for mental and behavioural disorders. After further adjustment for socioeconomic position and educational achievement (model 2), there was marked attenuation in all point estimates (range: 10.2-63.5%). Only three ADHD–health associations remained statistically significant: mental and behavioural disorders (3.51; 2.35, 5.22), diseases of the eye (2.04; 1.19, 3.33), diseases of the nervous system (1.81; 1.07, 2.91).

**Table 1.**
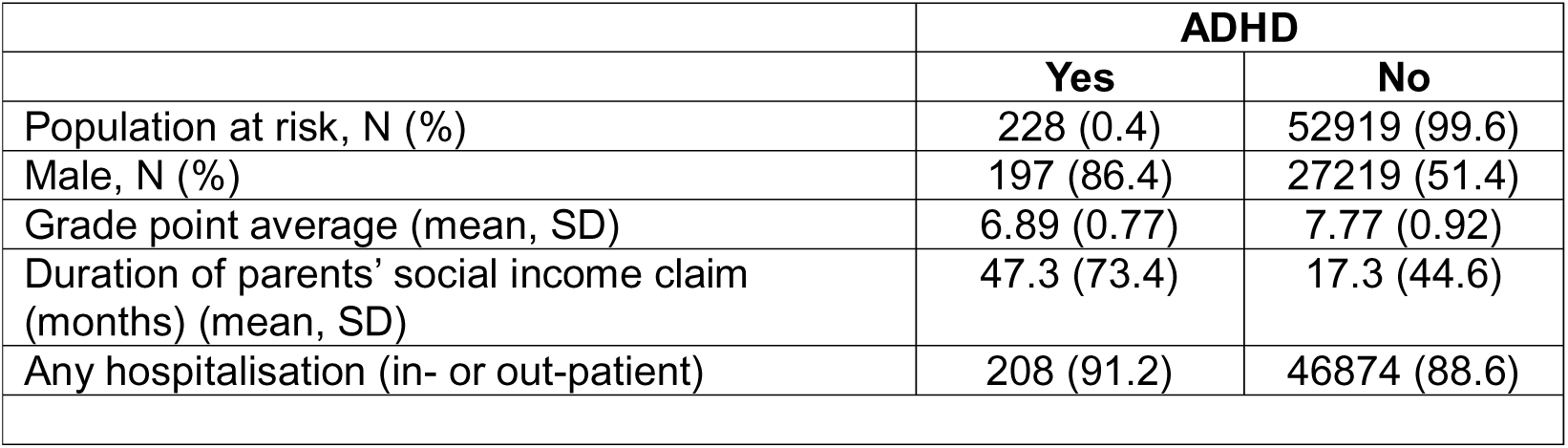
Study member characteristics according to ADHD diagnosis: The 1987 Finnish Birth Cohort (N=53147)

|  | **ADHD** |  |
| --- | --- | --- |
|  | **Yes** | **No** |
| Population at risk, N (%) | 228 (0.4) | 52919 (99.6) |
| Male, N (%) | 197 (86.4) | 27219 (51.4) |
| Grade point average (mean, SD) | 6.89 (0.77) | 7.77 (0.92) |
| Duration of parents' social income claim (months) (mean, SD) | 47.3 (73.4) | 17.3 (44.6) |
| Any hospitalisation (in- or out-patient) | 208 (91.2) | 46874 (88.6) |

Next, in figure 3 we show the association of ADHD diagnosis with 17 outcomes disaggregated from the ICD chapters in figure 2. Again, a higher risk of all these endpoints was seen in people with ADHD, however, only nine of these associations were statistically significant (99.7% confidence interval) after minimal adjustment and Bonferroni correction (model 1). After taking into account differences in parental socioeconomic status and study members’ education achievement, 5 associations remained: epilepsy (4.65; 1.86, 9.75), mood disorders (2.46; 1.50, 3.90), poisoning (2.30; 1.02, 4.51), substance abuse disorder (2.27; 1.28, 3.81), and neurotic disorders (2.12 [1.25−3.43]).

Lastly, to increase confidence in our new results for hospitalisations and antecedent ADHD, we attempted to replicate well-explored predictors of mortality in the same population (supplemental table 1). In sex-adjusted analyses, men (versus women odds ratio; 95% confidence interval: 2.95; 2.46, 3.56), study members with lower educational achievement (low versus high grade point average: 2.37; 2.01, 2.79), those from socioeconomically disadvantaged families (per SD longer duration of family income support: 1.28; 1.21, 1.34), and those diagnosed with ADHD (versus none: 2.15; 0.97, 4.08) all experienced a higher risk of premature mortality.

## Discussion

The main finding of the present study was that, after taking into account covariates and multiple testing in a phenotype-wide approach, pre-adult ADHD was associated with later occurrence of substance abuse, mood disorders, neurotic disorders, epilepsy, and poisoning. For those less well-examined health outcomes for which we had a working hypothesis – for example, bacterial/viral infection, road traffic accidents, and falls – while risk was significantly elevated in people with ADHD in basic analyses, there was marked attenuation to the point of non-significance after poverty and educational performance were added to the multivariable model.

### Comparison with existing studies

Our results add to the rapidly growing, if currently modest evidence base on the health consequences of ADHD. Consistent with other studies, we found a higher occurrence of mood disorders (depression),^13,14^ anxiety (neurotic) disorders,^15^ and substance abuse^16^ in people with ADHD relative to their unaffected peers. We did not have a sufficiently high number of people with substance use disorders to disaggregate this endpoint further. Meta-analyses suggest that, relative to population controls, people with ADHD are three times as likely to be nicotine-dependent^31^ and have a drug use disorder^32^ which includes marijuana^33^ and alcohol.^13^ Several psychological processes could explain the higher burden of these outcomes in the ADHD-affected, including self-medication whereby nicotine enhances mental focus and ethanol alleviates feelings of restlessness, hyperactivity and anxiety.^34^

We also found a raised risk of adult epilepsy in people diagnosed with ADHD in childhood/adolescence. Studies in this area have traditionally examined the role of epilepsy in the development of ADHD rather than the converse.^35^ In a rare exception, reports from a Taiwanese cohort also generated from registry linkages, suggest ADHD was associated with a four-fold elevation in the risk of this neurological condition in later life.^36^ While the pathways connecting epilepsy and ADHD have yet to be identified, there may be overlapping pathophysiologic and genetic mechanisms at play.

Lastly, in our study, people with ADHD had a higher likelihood of experiencing poisonings requiring hospital attention. Ascribing intention to episodes of poisonings is challenging for an attending-physician, however, those of deliberate^37^ and unknown aim^38^ have both be shown to be more common in people with ADHD in meta-analyses and large-scale administrative datasets, respectively. Deliberate poisoning accords with the literature on the elevated burden of self-harm and attempted suicide in people who are neurodiverse, particularly those with ADHD.^39^ When accidental, this could be a product of lower risk perception in people with an ADHD diagnosis who, on average, have lower cognitive function relative to the general population.^23^ This explanation has been advanced for the elevated risk of unintentional injury in people who achieve lower scores on standard IQ tests.^40^ In controlling for educational performance, a close correlated of IQ,^41^ the magnitude of the ADHD–poisoning relation, while clearly attenuated, was not eliminated. This would seem to implicate additional mechanisms underpinning this association.

### Study strengths and limitations

Our study has some strengths. These include the use of a full nation birth cohort with complete linkage to health records, so minimising concerns regarding selection bias and generalisability. Second, in our analyses we were able to replicate well-established findings in ADHD population-based research: a higher prevalence of being male, socioeconomically deprived, and diminished academic performance in individuals with a diagnosis of this neurodevelopmental condition. These characteristics are also known predictors of total mortality^42,43^ – again, observations made in the present dataset. Collectively, these findings give us confidence in the newer relationships reported here for ADHD and specific health conditions.

Our work is not without its limitations. First, all diagnoses – ADHD and health outcomes – were based on conditions severe enough to warrant hospital attention, whether in- or out-patient; people with less severe ADHD symptoms were therefore not captured. That the period prevalence of ADHD herein is based on administrative data may lead to some under-detection, such that studies which include sample-wide assessment of ADHD symptoms yield higher estimates.^10^ Relatedly, it is unclear if symptoms below current diagnostic thresholds for ADHD have an impact on adverse health events. Second, although we only computed effect estimates for those health endpoints with ten or more events, the breadth of the resulting confidence intervals suggests some analyses were underpowered. For instance, after disaggregating the outcomes in figure 2, confidence intervals in figure 3 are noticeably wider denoting lower precision. Third, in an attempt to ascertain independence of association, we adjusted for educational performance of the study member; this is a moot given that this characteristic may lie on mechanistic pathway connecting ADHD with injury and disease. Lastly, out-patient care episodes, including those for ADHD, were recorded from 1998 when study members were aged 11. As such, people with a prior ADHD diagnosis who did not continue to seek hospital care would not have been included in the ADHD group.

### Future research directions

ADHD, in contrast to other neurodevelopmental disorders such as autism and intellectual disability, is eminently treatable pharmacologically with psychostimulants, amongst other medications, and also psychosocial intervention which includes cognitive behavioural therapy.^44^ Well-controlled observational studies and clinical trials reveal that when core symptoms are successfully treated there is a concomitant positive impact on educational performance^45^ and quality of life,^46^ whereas discontinuation of therapy leads to a decline in grade point average.^47^ Whether treatment effects extend to the range of health outcomes associated with ADHD herein – as apparent for suicide attempt,^48^ unintentional injury,^22, 49^ and criminality^50^ – requires further study and we did not have data on prescriptions dispensed to investigate the impact of education.

While correlations between a growing array of adverse health outcomes and antecedent ADHD are being reported, these cohort studies, like our own, are generated using electronic health records. Such studies are typically not well-equipped to examine the mechanisms underpinning these associations.

For this to occur, field-based cohorts with higher resolution measurement of risk indices such as health behaviours (e.g., alcohol intake, physical activity) and biomarkers (e.g., inflammation, blood glucose) are required. Also, the impact of ADHD treatment, which has a significant effect on outcomes, was not included.

In conclusion, children and adolescents with ADHD in the present study had an increased future burden of psychological and neurological conditions but not somatic disorders.

## Data Availability

To discuss the availabilty of data, contact Mika Gissler

---

**Supplemental Table 1.** Study member characteristics in relation to total mortality in the 1987 Finnish Birth Cohort Study (N=59476)

|  | Total mortality |  |
| --- | --- | --- |
|  | N events / N at risk | OR (95% CI) |
| Male | 459 / 30435 | 2.95 [2.46-3.56] |
| Female | 150 / 29041 | 1.0 (ref) |
| Low grade point average | 294 / 14821 | 2.37 [2.01-2.79] |
| High grade point average | 315 / 44655 | 1.0 (ref) |
| Months claiming social income (per SD higher) | 609 / 59476 | 1.28 [1.21-1.34] |
| ADHD diagnosis | 8 / 279 | 2.15 [0.97-4.08] |
| ADHD free | 601 / 59197 | 1.0 (ref) |

ORs are sex-adjusted except in the analyses of risk according to sex where they are unadjusted. For GPA, ORs are for low (4-7) versus high [7-10]. Follow-up was to 2020. A SD increase in duration claiming social income is 45.74 months.

## Notes

Funding: The present paper received no direct funding. GDB is supported by the UK Medical Research Council (MR/P023444/1) and the US National Institute on Aging (1R56AG052519-01, 1R01AG052519-01A1).

### Competing Interest Statement

The authors have declared no competing interest.

### Funding Statement

The present paper received no direct funding. GDB is supported by the UK Medical Research Council (MR/P023444/1) and the US National Institute on Aging (1R56AG052519-01, 1R01AG052519-01A1).

### Author Declarations

Study approval was provided by the Ethical committee at the Finnish Institute for Health and Welfare (28/2009).

### Summary of Updates

Idiotically, I misspelt the name of one of my co-authors (Seyed Ehsan Mousavi). This has been corrected. There have been no other revisions.

